# Quantitative assessment of tumor immune microenvironment in context of HER2 expression in MIBC

**DOI:** 10.64898/2026.09.14.26363035

**Authors:** Wei Du, Shunsuke Koga, Ranjitha Pratap Nair, Rashmi Tondon, Paul J Zhang, Raghunath Puthiyaveettil, Lin Mei, Priti Lal

## Abstract

Muscle-invasive bladder cancer (MIBC) carries high risks of relapse and death despite contemporary care. In this study we aimed to evaluate whether the pretreatment immune microenvironment varied with HER2 status and if the differences influenced the response to neoadjuvant chemotherapy (NAC) and survival. We conducted a retrospective study of 62 treatment-naïve MIBC patients diagnosed on pretreatment transurethral resection of bladder tumor (TURBT) specimens collected between 2008 and 2024. All patients received NAC followed by radical cystectomy. HER2 immunohistochemistry (IHC) was scored 0/1+/2+/3+ using gastroesophageal criteria. PD-L1 was assessed by combined positive score (CPS). CD3, CD20, and CD68 cell densities were quantified by digital image analysis. Overall survival was measured from the date of TURBT. HER2 IHC scores were 2+/3+ in 28 of 62 tumors (45%) and 0/1+ in 34 (55%). All seven IHC 2+ tumors lacked ERBB2 amplification by fluorescence in situ hybridization (FISH). NAC response occurred in 11 of 34 tumors with HER2 IHC 0/1+ (32%) and 9 of 28 tumors with HER2 IHC 2+/3+ (32%) (P = 1.00). PD-L1 expression was not associated with HER2 status. Immune-cell analyses showed trends toward higher CD3+ T-cell density, CD20+ B-cell density, and TM ratio in NAC responders, but no significant differences by HER2 status or combined PD-L1/HER2 subgroup status. After Bonferroni correction for multiple comparisons, none of the immune-cell metrics remained statistically significant. Overall survival did not differ between the HER2 IHC groups (P = 0.71). These findings indicate that HER2 IHC 2+/3+ expression is present in a substantial subset of MIBC but is not associated with NAC response or overall survival in this cohort. Further study is warranted to define the clinical relevance of MIBC with HER2 IHC 2+/3+ and its potential role in HER2-directed therapeutic strategies.

## Introduction

Muscle-invasive bladder cancer (MIBC) is presented with one third of new bladder cancer cases each year. It is associated with high risks of relapse and death despite contemporary management.(1, 2) Neoadjuvant treatment followed by radical cystectomy improves survival; however, approximately 50% patients eventually experience metastasis or recurrence after surgery.(3) Identifying actionable molecular targets remains an unmet need in MIBC.

The human epidermal growth factor receptor 2 (HER2), encoded by *ERBB2* on chromosome 17q12, is a ligand-independent receptor tyrosine kinase of the ERBB family that promote proliferation and survival.(4) Since the introduction of HER2-targeted treatment revolutionized HER2-positive breast cancer, it has been extended later into gastrointestinal malignancies as well as non-small cell lung cancer, indicating HER2 as a validated therapeutic target in multiple solid tumors.(5–7) Antibody-drug conjugates have further expanded the scope of HER2 targeting to tumors with lower levels of expression HER2.(8) Trastuzumab deruxtecan (T-DXd) showed significant activity in progressive metastatic bladder cancer with in immunohistochemistry (IHC) 3+ disease.(9) Recently, phase 3 RC48-C016 trial showed that disitamab vedotin (DV) plus toripalimab improved progression-free and overall survival in HER2 expressing advanced urothelial cancer under the first line setting .(10) These data support HER2-directed therapy as a promising strategy in bladder cancer, though its efficacy in MIBC is sparse. Several challenges hinder further studies in MIBC. Notably, HER2 biology in urothelial carcinoma is heterogeneous.(11) Reported HER2 prevalence varies by assay and threshold.(12) Variability in scoring and intratumoral heterogeneity challenge interpretation, and there is no consensus yet of using gastric or breast cancer HER2 assessment criteria in the absence of urothelial-specific guidelines.

The tumor microenvironment is another determinant of outcome in MIBC. Prior studies suggest links between pre-treatment lymphocyte infiltration and response to neoadjuvant chemotherapy (NAC), including associations involving CD8 T cells and the balance of effector to regulatory compartments. In addition, PD-L1 expression using the combined positive score (CPS) is widely adopted in urothelial carcinoma as a strong prognostic marker.(13) However, the immune microenvironment and the association with HER2 expression is unknown.

Given recent gains with HER2-directed regimens in metastatic bladder cancer, we aimed to test whether HER2 status impacts immune microenviornment and outcomes of neoadjuvant cytotoxic chemotherapy. We analyzed a single-center retrospective cohort of 62 treatment-naïve MIBC cases diagnosed on pre-treatment transurethral resection of bladder tumor (TURBT). HER2 was scored by IHC using gastroesophageal criteria,(14) PD-L1 was assessed by CPS, and CD3, CD20, and CD68 densities were quantified as cells/mm². We examined associations with clinicopathologic features, pathologic downstaging after NAC, and overall survival.

## Materials and Methods

### Study design and cohort

This retrospective, single-center cohort comprised 62 consecutive patients with MIBC. The index pretreatment TURBT specimens analyzed in this study were collected between 2008 and 2024 at the Hospital of the University of Pennsylvania. Inclusion criteria required identification of patients who underwent in-house index TURBT procedure, with a MIBC and subsequently received NAC followed by radical cystectomy at our institution. Exclusion criteria were prior resection at an outside institution or previous local or systemic therapy for bladder cancer. All hematoxylin and eosin-stained slides were reviewed by board-certified genitourinary pathologists to confirm diagnosis, tumor content, and histologic variant. One representative slide and its corresponding formalin fixed paraffin embedded tissue block was selected for each case. All slides were scanned at 20x magnification using a HAMAMATSU NanoZoomer S360 scanner. Digital slide visualization and image analysis were performed using QuPath, an open-source software platform for digital pathology image analysis. The primary endpoint was NAC response defined by pathologic downstaging on cystectomy relative to pretreatment stage. Overall survival was measured from the date of TURBT to death or last contact. This study was conducted in accordance with the Declaration of Helsinki and was approved by the Institutional Review Board (IRB) of the Hospital of the University of Pennsylvania. The requirement for informed consent was waived by the Institutional Review Board.

A subset of this cohort was included in a study presented at the 2025 United States and Canadian Academy of Pathology (USCAP) Annual Meeting.(15) Preliminary findings from the present study were presented at the 2026 USCAP Annual Meeting.(16)

### Clinical and Pathology Data Collection

Clinical and pathological variables were abstracted from the electronic medical record. These included age at TURBT, sex, and race, tumor size and volume in the TURBT specimen, histologic subtype, and clinical stage. Treatment data included NAC regimen, number of cycles, and start and stop dates. From cystectomy reports, we recorded the procedure type, post-NAC pathologic stage at cystectomy, greatest tumor dimension, lymphovascular invasion when present, and margin status. Dates of first metastasis, last follow-up, and death were obtained from oncology and urology documentation. Two investigators performed independent data checks, and discrepancies were resolved by consensus before analysis.

### Immunohistochemical Staining

Formalin-fixed, paraffin-embedded sections from pre-treatment TURBT specimens were stained for HER2, PD-L1, CD3, CD20, and CD68. IHC for HER2 was performed using the fully automated Ventana BenchMark ULTRA platform and the UltraView Universal DAB detection system (Ventana 760-500). HER2 expression was assessed with the prediluted PATHWAY anti-HER2/neu (4B5) monoclonal antibody (Ventana, 790-100) after heat-induced epitope retrieval with Cell Conditioner 1 (CC1; Ventana, 950-224) for 36 minutes, according to the manufacturer’s instruction.

PD-L1 IHC was performed on BenchMark ULTRA platform using the OptView DAB IHC detection kit (Ventana 760-500). Sections were pretreated with CC1 (Ventana, 950-224) for 64 minutes and then incubated with an anti–PD-L1 monoclonal antibody (Dako, M365329-1; clone 22C3; 1:50), according to the manufacturer’s instruction.

IHC for CD3, CD20, and CD68 was performed on a Leica BOND-PRIME instrument with the Bond Prime Polymer Refine Detection System (Leica, AR0087). Heat-induced epitope retrieval was carried out on-instrument using Bond Prime Epitope Retrieval Solution 2 (ER2; Leica, AR0087) for 20 minutes for CD3 and CD20, and Bond Prime Epitope Retrieval Solution 1 (ER1; Leica, AR0086) for 20 minutes for CD68. After epitope retrieval, prediluted primary antibodies were applied as follows: CD3 (Leica, PA0553; clone L10), CD20 (Dako, IR60461; clone L26), and CD68 (Dako, IR609; clone KP1). Primary antibody incubation and subsequent post-primary and polymer steps were executed on-instrument using the Bond Prime Polymer Refine protocol according to the manufacturer’s instructions. Sections were counterstained with hematoxylin and mounted for microscopic examination.

### Immunostaining score and digital image analysis

HER2 IHC was scored 0, 1+, 2+, or 3+ according to standardized gastroesophageal criteria.(14) Score 3+ was defined as strong complete or basolateral membranous staining in ≥10% of invasive tumor cells. Score 2+ was defined as weak to moderate complete or basolateral membranous staining in ≥10% of cells. Score 1+ was defined as faint, incomplete membranous staining in ≥10% of cells. Score 0 was defined as no membranous staining or staining in <10% of cells. Representative examples of scores 0, 1+, 2+, and 3+ are shown in **Figure 1**. For statistical analyses, tumors were grouped by HER2 IHC score as 0/1+ versus 2+/3+.

**Figure 1:**
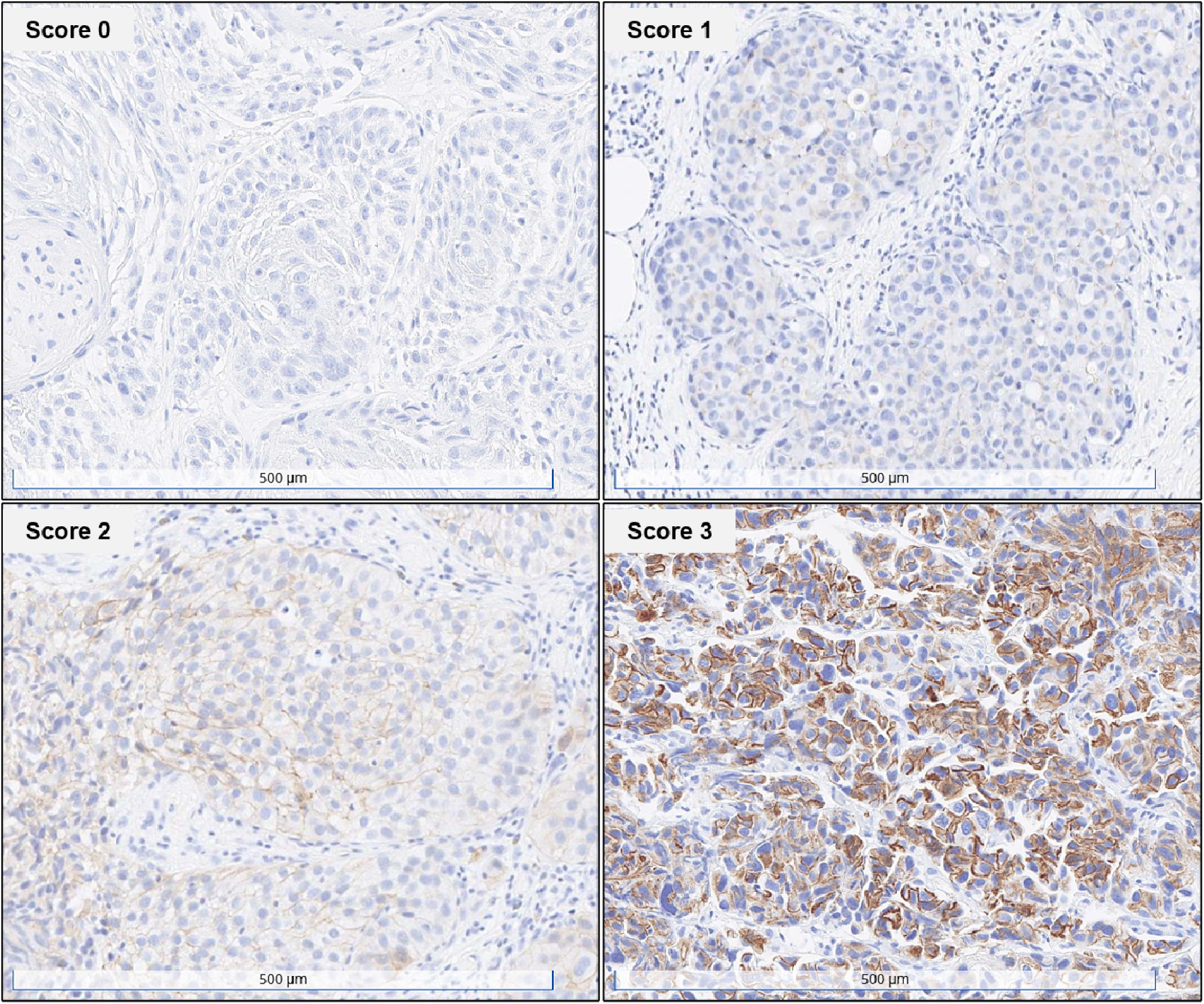
HER2 immunohistochemistry on pre-treatment TURBT. Representative images show HER2 IHC scores 0, 1+, 2+, and 3+ in muscle invasive bladder cancer.

PD-L1 was recorded as the CPS, defined as 100 × (number of PD-L1–positive tumor cells, lymphocytes, and macrophages) divided by the number of viable tumor cells, capped at 100.(13) For reporting, we summarized the proportion of PD-L1–positive tumors using a prespecified cutoff of CPS ≥1 and recorded CPS as a continuous variable for secondary analyses, consistent with prior urothelial carcinoma trials that used CPS.(17)

For quantitative immune profiling, whole-slide images were generated by scanning IHC slides for CD3, CD20, and CD68 at 20× magnification. Images were analyzed in QuPath.(18) Tumor-rich regions of interest were manually annotated by a pathologist, and areas with cautery artifact, necrosis, crush change, or non-neoplastic tissue were excluded. Positive cell detection was performed after hematoxylin and DAB color deconvolution using standardized thresholds optimized on a training subset. For each case, positive-cell counts were normalized to annotated tumor area and expressed as cell density (cells/mm²). A T cell-to-macrophage ratio (TM ratio) was calculated for each case as the CD3+ cell density divided by the CD68+ cell density.

### Fluorescence in situ hybridization

ERBB2 amplification was assessed by fluorescence in situ hybridization (FISH) in all seven tumors with a HER2 IHC score of 2+. FISH was performed in-house on 4–5-μm sections of formalin-fixed, paraffin-embedded tissue using the PathVysion HER2 DNA Probe Kit (Vysis). The dual-color assay targeted HER2 and the chromosome 17 centromere (CEP17). Following deparaffinization and pretreatment, hybridization and post-hybridization washing were performed according to the laboratory protocol.

Nuclei were counterstained with DAPI. Tumor areas were identified using a corresponding hematoxylin and eosin-stained section from the same block. HER2 and CEP17 signals were enumerated in 30–80 evaluable interphase tumor nuclei per case. Mean HER2 and CEP17 copy numbers and the HER2/CEP17 ratio were calculated. A positive control was included in each assay run.

FISH results were reviewed according to the CAP/ASCP/ASCO guidelines for gastroesophageal adenocarcinoma.(14) Interpretation incorporated the HER2/CEP17 ratio and the mean HER2 and CEP17 copy numbers. FISH findings were used to characterize ERBB2 amplification. For association analyses, tumors were grouped as HER2 IHC 0/1+ or 2+/3+, irrespective of FISH results.

### Statistical Analysis

Descriptive statistics summarize demographic, pathological, and biomarker variables. Histologic subtype distributions were compared using the Fisher–Freeman–Halton exact test. Other categorical variables were analyzed using chi-squared tests. CD3+, CD20+, and CD68+ cell densities and TM ratio were compared between NAC responders and non-responders, and separately between tumors with HER2 IHC scores of 2+/3+ and 0/1+, using the Mann-Whitney U test. Four-group comparisons stratified by combined PD-L1 and HER2 status were performed using the Kruskal-Wallis test. All P values were two-sided. Bonferroni correction was applied across the 12 comparisons of immune-cell density and TM ratio, with P < 0.00417 considered statistically significant. Analyses and graphics were generated using GraphPad Prism (Graphpad Holdings, LLC) and Python. Kaplan-Meier survival analysis and log-rank testing were performed in Python, and survival curves were plotted using the lifelines and matplotlib packages.

## Results

### NAC response cohort

Baseline characteristics are summarized in **Table 1**. The cohort included 62 patients, of whom 20 were responders and 42 were non-responders based on cystectomy restaging. All patients had clinical stage II or III disease, including 38 (61.3%) with stage II disease. Stage II disease was more frequent among responders than nonresponders (85% vs 50%, P = 0.008). Median age at TURBT was similar between groups (65 [62, 71] vs 69 [64, 76] years). Men comprised 75% of responders and 57% of non-responders. Caucasian race was recorded as 75% and 79%, respectively. Most patients received cisplatin/gemcitabine as NAC (responders 17/20, 85%; nonresponders 23/42, 55%). Additional regimens included cisplatin/gemcitabine plus pembrolizumab (0% vs 14%), and other regimens (10% vs 29%). NAC regimen details were unavailable for two patients, one in each response group.

**Table 1:** Baseline demographic, clinical, and pathologic characteristics by neoadjuvant chemotherapy response.

|  | **Responder**<br>**(n = 20)** | **Non-Responder**<br>**(n = 42)** | **P value** |
| --- | --- | --- | --- |
| **Demographics** |  |  |  |
| Sex, %Male | 15 (75%) | 24 (57%) | 0.17 |
| Race, %Caucasian | 15 (75%) | 33 (79%) | 0.75 |
| Initial clinical stage (stage II) | 17 (85%) | 21 (50%) | 0.01 |
| **Clinical course** |  |  |  |
| Age at TURBT,* years | 65 [62, 71] | 69 [64, 76] | 0.10 |
| Follow-up duration,* month | 60 [54, 77] | 13 [5, 26] | **<0.001** |
| Metastasis during follow-up | 2 (10%) | 31 (74%) | **<0.001** |
| Death during follow-up | 5/20 (25%) | 33/41 (81%) | **<0.001** |
| **Chemotherapy** |  |  | 0.07 |
| Cisplatin/Gemcitabine | 17 (85%) | 23 (55%) |  |
| Cisplatin/Gemcitabine/Pembrolizumab | 0 (0%) | 6 (14%) |  |
| Other regimens | 2 (10%) | 12 (29%) |  |
| Not available | 1 (5%) | 1 (2%) |  |
| **Pathology subtype** |  |  | 0.161 |
| Urothelial carcinoma, NOS | 8 (40%) | 24 (57%) |  |
| Micropapillary | 2 (10%) | 3 (7%) |  |
| Sarcomatoid | 0 (0%) | 5 (12%) |  |
| Squamous | 7 (35%) | 6 (14%) |  |
| Other | 3 (15%) | 4 (10%) |  |
| Tumor volume,* cm<sup>3</sup> | 14.0 [4.4, 29.7] | 12.6 [4.8, 34.3] | 0.74 |
| Cystectomy tumor size, greatest dimension, cm | 0.0 [0.0, 0.1] | 4.0 [2.1, 5.5] | **<0.001** |

\*Data are shown median [25%tile, 75%tile] and analyzed by Mann–Whitney U test. Histologic subtype distributions are compared using the Fisher–Freeman–Halton exact test. Other categorical data are analyzed using the chi-squared test. Percentages and P values for death during follow-up are calculated among patients with known vital status (n = 61). Abbreviations: TURBT, transurethral resection of bladder tumor; NOS, not otherwise specified.

Histologic variants on TURBT were not different between two groups. Baseline tumor volume on TURBT did not differ (14.0 [4.4, 29.7] vs 12.6 [4.8, 34.3] cm³) between responders and non-responders.

Responders had longer follow-up (60 [54, 77] vs 13 [5, 26] months) and a lower rate of metastasis. Among patients with known vital status, death during follow-up occurred in 5 of 20 responders (25.0%) and 33 of 41 nonresponders (80.5%) (P < 0.001).. Post NAC-Cystectomy tumor size was smaller in responders, consistent with pathological downstaging.

### HER2 and PD-L1 expression

HER2 IHC scores in pretreatment TURBT specimens were 3+ in 21/62 tumors (33.9%), 2+ in 7/62 (11.3%), and 0/1+ in 34/62 (54.8%). All seven tumors with HER2 IHC 2+ were classified as negative for HER2 amplification according to the institutional FISH interpretation criteria. HER2/CEP17 ratios ranged from 1.27 to 1.98, and mean HER2 copy numbers ranged from 2.57 to 5.24 per cell.

For group comparisons, tumors were classified as HER2 IHC 0/1+ (n = 34) or 2+/3+ (n = 28), irrespective of FISH results. Clinicopathological characteristics by HER2 IHC status are summarized in **Table 2**. There are no differences between sex, races, median age, chemotherapy regimens and outcomes. The distribution of pathological subtypes, including urothelial carcinoma NOS, micropapillary, sarcomatoid, squamous and others, are similar between HER2 IHC 2+/3+ and 0/1+ groups.

**Table 2:** Baseline demographic, clinical, and pathologic characteristics by HER2 status.

|  | HER2-Negative<br>(n = 34) | HER2-Positive<br>(n = 28) | P value |
| --- | --- | --- | --- |
| **Demographics** |  |  |  |
| Sex, %Male | 21 (62%) | 18 (64%) | 0.84 |
| Race, %Caucasian | 29 (85%) | 19 (68%) | 0.13 |
| Initial clinical stage (stage II) | 21 (62%) | 17 (61%) | 0.93 |
| **Clinical course** |  |  |  |
| Age at TURBT,* year | 69.5 [63.2, 76.0] | 66.5 [62.0, 71.8] | 0.17 |
| Follow-up duration, months | 20.5 [5.25, 48.5] | 20.5 [6.0, 62.0] | 0.60 |
| Recurrence during follow-up | 17 (50%) | 19 (68%) | 0.16 |
| Death during follow-up | 19/33 (58%) | 19/28 (68%) | 0.41 |
| Overall survival, months | 40.2 [12.2, 61.5] | 31.9 [13.4, 70.6] | 0.93 |
| **Pathology subtype** |  |  | 0.50 |
| Urothelial carcinoma, NOS, n (%) | 15 (44%) | 17 (61%) |  |
| Micropapillary, n (%) | 2 (6%) | 3 (11%) |  |
| Sarcomatoid, n (%) | 4 (12%) | 1 (4%) |  |
| Squamous, n (%) | 9 (26%) | 4 (14%) |  |
| Other, n (%) | 4 (12%) | 3 (11%) |  |
| Tumor volume,* cm<sup>3</sup> | 11.5 [4.3, 38.2] | 13.8 [5.3, 27.3] | 0.73 |
| Cystectomy tumor size, greatest dimension, cm | 3.1 [0.1, 5.0] | 2.1 [0.5, 3.8] | 0.41 |

\*Data are shown median [25%tile, 75%tile] and analyzed by Mann–Whitney U test. Histologic subtype distributions are compared using the Fisher–Freeman–Halton exact test. Other categorical data are analyzed using the chi-squared test. Percentages and P values for death during follow-up are calculated among patients with known vital status (n = 61). Abbreviations: TURBT, transurethral resection of bladder tumor; NOS, not otherwise specified.

PD-L1 was assessed in 62 tumors and was positive in 17 (27%) and negative in 45 (73%) cases. Among tumors with HER2 IHC 0/1+ (n = 34), 11 (32.4%) were PD-L1 positive and 23 (67.6%) were negative. Among tumors with HER2 IHC 2+/3+ (n=28), 6 (21.4%) were PD-L1 positive and 22 (78.6%) were negative. The mean PD-L1 combined positive score (CPS) was 5.18 ± 11.8. PD-L1 positivity did not differ significantly between the HER2 IHC 0/1+ and 2+/3+ groups (P = 0.50).

### HER2 status and pathological response to NAC

HER2 IHC scores stratified by pathological response to NAC are summarized in **Table 3**. Among responders, HER2 IHC was 3+ in 5 of 20 tumors (25.0%), 2+ in 4 (20.0%), 1+ in 0, and 0 in 11 (55.0%). Among non-responders, HER2 IHC was 3+ in 16 of 42 tumors (38.1%), 2+ in 3 (7.1%), 1+ in 2 (4.8%), and 0 in 21 (50.0%). Pathological response occurred in 11 of 34 tumors with HER2 IHC 0/1+ (32.4%) and in 9 of 28 tumors with HER2 IHC 2+/3+ (32.1%). The response rates did not differ significantly between the groups (P = 1.00).

**Table 3:**
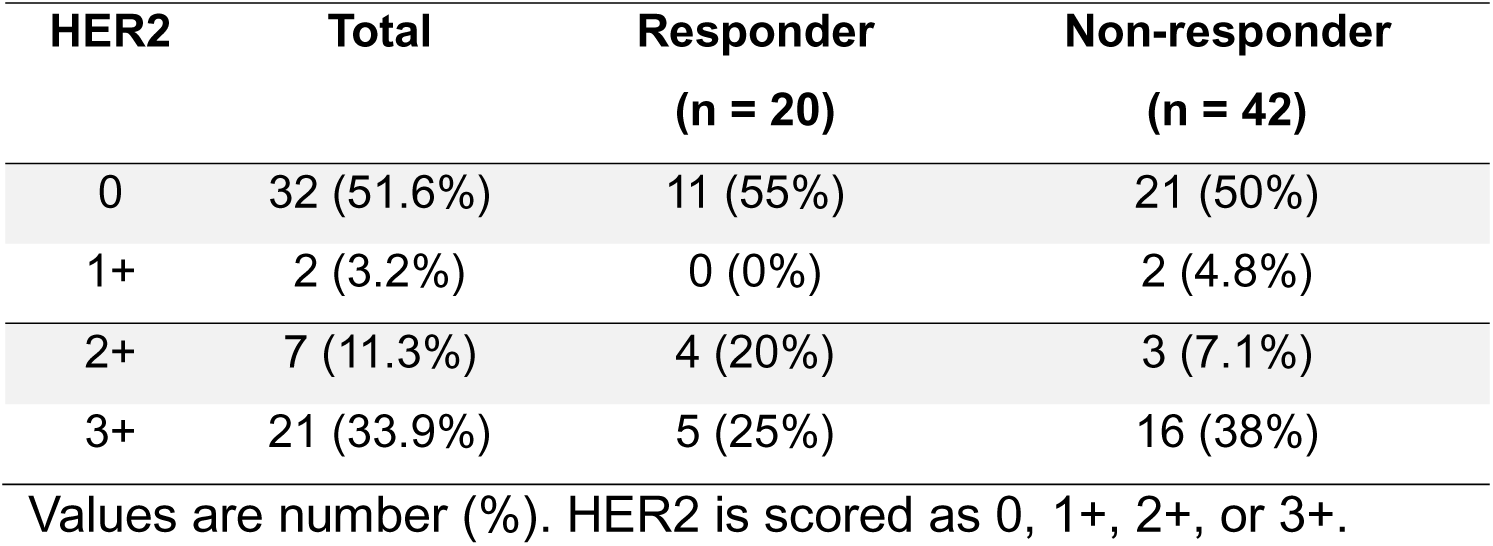
Distribution of HER2 immunohistochemistry scores overall and by neoadjuvant chemotherapy response.

### Tumor immune microenvironment and chemotherapy response

Quantitative immune profiling of pretreatment TURBT showed trends toward an inflamed microenvironment in NAC responders. CD3+ T-cell density, CD20+ B-cell density, and the TM ratio were higher in responders than in non-responders, whereas CD68+ macrophage density did not differ between groups (**Figure 2**, top panels). When tumors were analyzed by HER2 IHC status (2+/3+ vs 0/1+), no significant differences were observed in CD3+, CD20+, or CD68+ cell densities or in TM ratio (Figure 2, middle panels). Further subgroup analyses stratified by combined PD-L1 and HER2 status also showed no significant differences in immune-cell densities or TM ratio (Figure 2, bottom panels). After Bonferroni correction for multiple comparisons, none of these associations remained statistically significant.

**Figure 2.**
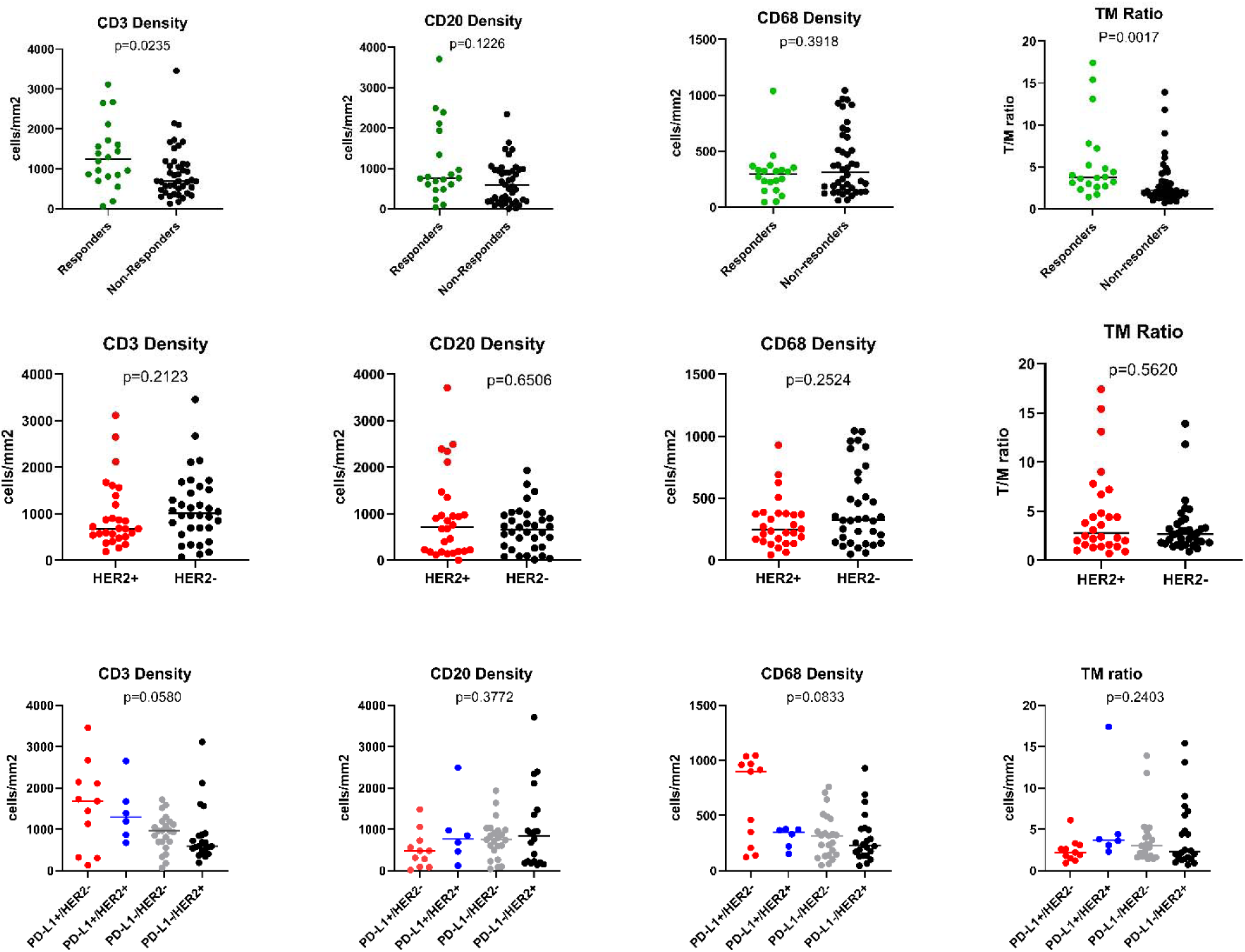
Immune-cell densities according to NAC response, HER2 IHC status, and combined PD-L1/HER2 subgroup status. Dot plots show CD3+, CD20+, and CD68+ cell densities (cells/mm²) and TM ratio quantified on pre-treatment TURBT specimens using QuPath. The top row shows comparisons between NAC responders and non-responders. The middle row shows comparisons between tumors with HER2 IHC scores of 2+/3+ and 0/1+. The bottom row shows four-group comparisons stratified by combined PD-L1 and HER2 status: PD-L1-negative/HER2 0/1+ (n = 23), PD-L1-positive/HER2 0/1+ (n = 11), PD-L1-negative/HER2 2+/3+ (n = 22), and PD-L1-positive/HER2 2+/3+ (n = 6). P values are shown in each panel. The Mann-Whitney U test is used for the top and middle rows, and the Kruskal-Wallis test is used for the bottom row. Bonferroni correction is applied across the 12 comparisons, with P < 0.00417 considered statistically significant.

### Clinical outcomes and survival

Follow-up was longer in responders than in non-responders (median 60 [54, 77] vs 13 [5, 26] months; P < 0.001). As expected, metastasis during follow-up developed in 2 of 20 responders (10%) and in 31 of 42 non-responders (74%) (P < 0.001). Death occurred in 5 of 20 responders (25%) and in 33 of 41 non-responders (81%) (P < 0.001). The proportion of deaths did not differ significantly between the HER2 IHC 0/1+ and 2+/3+ groups (19/33 [57.6%] vs 19/28 [67.9%], P = 0.41). Residual tumor at cystectomy was smaller in responders than in non-responders (0.0 [0.0, 0.1] vs 4.0 [2.1, 5.5] cm; P < 0.001), consistent with effective downstaging. In Kaplan-Meier analysis from TURBT (**Figure 3**), overall survival was similar between tumors with HER2 IHC 2+/3+ (n = 28) and 0/1+ (n = 34), and the log-rank test was not significant (P = 0.71).

**Figure 3.**
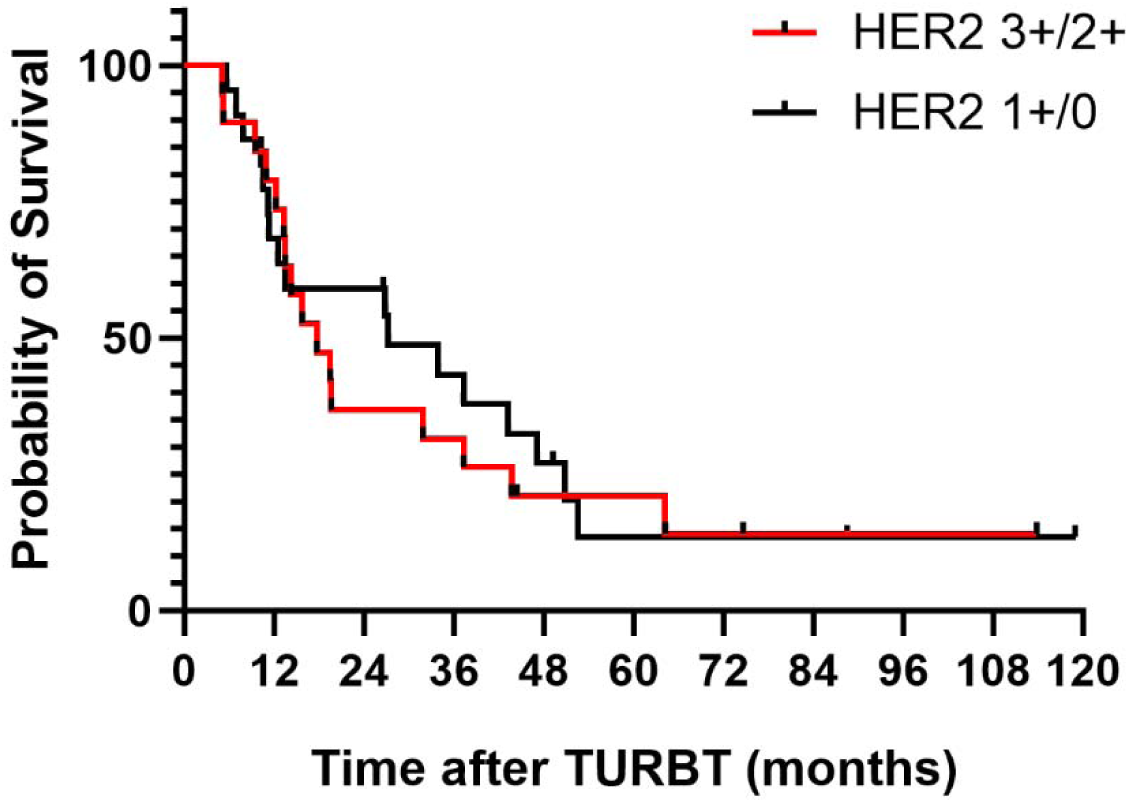
Overall survival after TURBT by HER2 IHC status. Kaplan-Meier curves show overall survival from TURBT stratified by HER2 IHC status (2+/3+ vs 0/1+). The difference is not statistically significant (log-rank P = 0.71).

## Discussion

In this single-institution study of pre-treatment TURBT specimens from patients with MIBC, approximately one third of tumors showed HER2 IHC 3+, and 45% were IHC 2+ or 3+. These frequencies are consistent with reports in metastatic urothelial carcinoma when comparable definitions and assays are applied, although published estimates vary widely across studies.(10, 11, 19) Because a urothelial-specific standard for HER2 IHC interpretation in MIBC has not yet been established, we adopted the ASCO/CAP gastroesophageal adenocarcinoma criteria for scoring to improve reproducibility and comparability.(14) The enrichment of HER2 expression in micropapillary and other aggressive variants is consistent with genomic studies that demonstrate frequent *ERBB2* amplification or mutation in micropapillary carcinoma.(12, 20, 21) FISH was performed to further characterize ERBB2 amplification in the seven IHC 2+ tumors, all of which were reported as nonamplified by the testing laboratory. Disitamab vedotin has shown activity in metastatic urothelial carcinoma with HER2 IHC 2+ or 3+, including IHC 2+/FISH-negative tumors.(22) This provides a clinical rationale for our IHC 0/1+ versus 2+/3+ grouping, which reflects protein expression irrespective of ERBB2 amplification.

In our cohort, HER2 status was not associated with NAC response, PD-L1 CPS, or overall survival. These findings do not support a clear predictive or prognostic role for HER2 in the current treatment setting of MIBC. This is in line with prior studies in advanced urothelial carcinoma suggesting that ERBB2 alteration status does not necessarily predict outcome with standard systemic therapy.(23) Our data extend this observation to the neoadjuvant setting by showing that HER2 IHC status was not associated with pathologic response to NAC.

Response to HER2-directed therapy may vary with the level and biology of HER2 expression. In the bladder cancer cohort of DESTINY-PanTumor02, objective response rates with T-DXd were 56.3% and 35.0% in tumors with centrally assessed HER2 IHC 3+ and 2+, respectively.(9) In 2024, the FDA granted accelerated approval to T-DXd for previously treated adults with unresectable or metastatic IHC 3+ solid tumors who lack satisfactory alternative treatment options. The phase III RC48-C016 trial demonstrated improved progression-free and overall survival with disitamab vedotin plus toripalimab compared with chemotherapy in advanced urothelial carcinoma with HER2 IHC 1+, 2+, or 3+.(10) HERALD/EPOC1806 basket trial selected patients with advanced solid tumors based on ERBB2 amplification detected by plasma cell-free DNA testing, without requiring HER2 IHC for enrollment. The independently assessed objective response rate with T-DXd was 58.1%.(24) These observations highlight the need for standardized HER2 assessment in bladder cancer, potentially incorporating ISH in equivocal cases. Our findings indicate that such testing is feasible in MIBC using TURBT specimens.

The tumor microenvironment may also contribute to treatment response in MIBC. Prior studies have suggested that increased pretreatment lymphocytic infiltration is associated with chemosensitivity and favorable outcomes in MIBC.(25, 26) B cell-rich tertiary lymphoid structures have also been linked to improved survival and immunotherapy response in bladder cancer.(27) In our cohort, pretreatment TURBT specimens from NAC responders showed trends toward higher CD3+ T-cell density, CD20+ B-cell density, and TM ratio. However, these associations did not remain statistically significant after Bonferroni correction for multiple comparisons. In addition, exploratory subgroup analyses based on combined PD-L1 and HER2 status showed no significant differences in immune-cell densities or TM ratio. These findings suggest that immune contexture may be relevant to treatment sensitivity, but the current dataset is insufficient to support conclusions regarding biomarker-defined immune subsets.

This study has limitations. It is retrospective and from a single center, and the sample size was modest, particularly for histologic variants and biomarker-defined subgroup analyses. HER2 IHC was scored using gastroesophageal criteria because bladder-specific guidelines are not established, and assay choice may influence positivity rates.(28) Because FISH was restricted to IHC 2+ tumors, the prevalence and prognostic significance of ERBB2 amplification in the overall cohort were not evaluated. Immune-cell quantification was performed in selected immune-rich regions of TURBT specimens and normalized to tissue area. These measurements may not fully represent spatial heterogeneity across the tumor. Particularly, anti-HER2 treatment was not utilized in this cohort. Despite these limitations, the study benefits from uniform pre-treatment specimens, centralized pathology review, standardized HER2 scoring, and quantitative digital pathology.

In conclusion, HER2 IHC 2+/3+ expression was observed in a substantial subset of pretreatment MIBC, but HER2 status was not associated with NAC response, PD-L1 CPS, or overall survival in this cohort. Immune-cell analyses suggested differences in pretreatment tumor microenvironment by NAC response, but these associations did not remain significant after multiple-comparison correction. Exploratory analyses of combined PD-L1/HER2-defined subgroups were limited by sample size and showed no significant differences. These findings support further evaluation of MIBC with HER2 IHC 2+/3+ as a candidate population for HER2-directed therapy and provide a rationale for prospective studies with standardized biomarker assessment and planned tissue-based correlative analyses.

## Acknowledgment

We thank the pathology assistants and residents in the Department of Pathology and Laboratory Medicine at the Hospital of the University of Pennsylvania for assistance with specimen preparation. We also thank Li-Ping Wang, Research Specialist at the Pathology Clinic Service Center, for performing the immunohistochemical staining for this study.

## Funding Statement

This study was supported in part by a Resident Research Grant from the Department of Pathology and Laboratory Medicine at the Hospital of the University of Pennsylvania (to Wei Du) and by the University of Pennsylvania McCabe Fellow Award (to Lin Mei).

## Competing Interests

The authors declare no competing interests.

## Data Availability

The data supporting the findings of this study are not publicly available because they contain sensitive patient information. Requests for access to de-identified data should be directed to the corresponding authors and will be considered subject to institutional policies and applicable approvals.

## Author Contributions

Wei Du: data collection; visualization; writing (review); funding acquisition. Shunsuke Koga: data collection; formal analysis; visualization; writing (original draft). Ranjitha Pratap Nair: assistance with data collection; writing (review). Rashmi Tondon: pathology evaluation; writing (review). Paul J. Zhang: FISH evaluation; Raghunath Puthiyaveettil: FISH evaluation; Lin Mei: data collection; writing (review and editing); funding acquisition. Priti Lal: pathology evaluation and diagnosis; writing (review and editing).

## Notes

### Competing Interest Statement

The authors have declared no competing interest.

### Author Declarations

This study was conducted in accordance with the Declaration of Helsinki and was approved by the Institutional Review Board (IRB) of the Hospital of the University of Pennsylvania. The requirement for informed consent was waived by the Institutional Review Board.

